# Diagnostic accuracy and predictive value of clinical symptoms for the diagnosis of mild COVID-19

**DOI:** 10.1101/2021.03.05.21252963

**Authors:** V. Popovych, I. Koshel, Y. Haman, V. Leschak, R. Duplikhin

## Abstract

**Objective:** To assess the diagnostic accuracy of clinical symptoms and their predictive values in patients with suspected mild COVID-19 and to identify target groups for self-isolation and outpatient treatment without additional testing in the primary health care system.

**Methods:** We conducted an open-label prospective study in both male and female patients aged 18 to 72 years with suspected mild COVID-19 who were sequentially enrolled in the study. The clinical diagnosis was performed in accordance with the WHO recommendations based on the acute onset of such symptoms as olfactory dysfunction, hyperthermia, myalgia, nasal congestion, nasal discharge, cough, rhinolalia, sore throat, without pneumonia or hypoxia in persons in contact with a confirmed case of COVID-19. The physician assessed clinical symptoms using a 4-point scale. The patient self-assessed clinical symptoms using a ten-point visual analogue scale (VAS). All enrolled patients underwent laboratory testing to confirm the diagnosis of COVID-19.

**Results:** Of the 120 patients underwent testing, the diagnosis of mild COVID-19 was confirmed in 96 patients and ruled out in 24 patients. When assessing symptoms by a physician according to the correlation analysis, hyperthermia, myalgia, nasal congestion and rhinolalia have a positive predictive value with a significance level of more than 0.6. When self-assessing symptoms by a patient, fever, myalgia and nasal congestion have a diagnostic accuracy with a significance level of more than 0.5. Nasal discharge, cough and sore throat have negative predictive values.

**Discussion:** The presence of these symptoms in patients with an acute onset of the disease can help to make a clinical diagnosis of coronavirus disease and identify target groups for self-isolation and outpatient treatment without additional testing. Highly suspect asymptomatic patients are not considered as those who have possible mild COVID-19 infection.

**Registration:** Ethics Committee of Ivano-Frankivsk National Medical University, Protocol No. 114/20 as of 21 May 2020.

## 1. Introduction

The novel 2019-nCoV coronavirus is of great concern since the virus is rapidly spreading around the world and the number of cases and deaths continues to rise. The main cause of mortality is cases of severe COVID-19, which include pneumonia, acute respiratory distress syndrome (ARDS), sepsis and septic shock [1]. Severe symptoms which require hospitalization are observed in about 20% of patients, and about 3% of reported cases require intensive care [2, 3]. Severe course and death are more common among the elderly or patients with comorbid chronic conditions [4].

According to updated data, 40% to 80% of COVID-19 patients have a mild disease [5,6]. At the same time, mild or asymptomatic cases are of great danger in terms of the spread of infection since the main source of infection is a sick person. The basic reproduction number (R_0_ is the average number of secondary cases generated by a primary case) of SARS-CoV-2 is 1.4 to 6.47. Thus, the patient even with mild manifestations may infect from 1.5 to 3.5 people and spread the virus to approximately 368 people in just five cycles of infection [7,8].

In view of the novelty of the virus, it is reasonable to assume that no one has immunity; that is, the entire human population is potentially susceptible to SARS - CoV-2 infection [9]. Modelling shows that paucisymptomatic infections can change the epidemic proportions and mortality rate. The SARS-CoV model predicts that there will be many more paucisymptomatic or unreported cases at the peak of infections which suggests that the widespread community transmission may occur [10]. This is supported by a modelling study on transmission in China, which suggests that about 86% of infections were due to unreported cases [11]. Studies show that paucisymptomatic patients are a spreading factor of SARS-CoV-2 and should be taken into account when predicting the epidemic proportions and the effectiveness of mitigation strategies. In its updated strategies to prevent the spread of COVID-19, WHO expresses the need for diagnosis and effective treatment of patients with a mild and moderate disease [12].

Mild COVID-19 is characterized by well-known non-specific manifestations: fever, cough, sore throat, nasal congestion, malaise, headache and muscle pain [13]. These symptoms are typical for non-severe acute viral upper respiratory tract infections, in particular acute nasopharyngitis, caused by already known human coronaviruses, in particular hCoV-229E, OC43, NL63 and HKU1 [14]. It has been proven that olfactory dysfunction is considered as a clinical marker for COVID-19 under pandemic conditions [15,16]. In Germany, 2 of 3 confirmed cases are reported to have anosmia [17]. These patients need to be verified as they can be hidden carriers who contribute to the rapid spread of COVID-19.

However, according to Chinese data, only 44,672 (62%) of 72,314 cases were classified as confirmed cases of COVID-19 and 16,186 (22%) cases — as suspicious. In these cases, no testing was performed since the testing capacity was insufficient to meet current needs. There were 10,567 (15%) clinically diagnosed cases. In these cases, no testing was performed and the diagnosis was made based on symptoms, contacts and signs on the lung imaging specific to coronavirus pneumonia. Eight hundred and eighty-nine (889) cases were asymptomatic (1%). The diagnosis was confirmed by a viral nucleic acid test but without typical symptoms including fever, dry cough and fatigue. The mild form was recorded in 81%, severe — in 14%, critical — in 5% [18].

Thus, almost 38% of patients were diagnosed with COVID-19 based only on clinical data, including epidemiological, laboratory and radiological data. At the same time, we still have insufficient information about the real course of mild disease [19,20]. Only acute onset of the disease and olfactory dysfunction are considered symptoms which require serious consideration for self-isolation and testing of patients [21,22]. Under present circumstances of the total spread of the pandemic and still limited testing capacity, there are no clear criteria to make a clinical diagnosis of mild COVID-19 and to identify individuals for self-isolation and outpatient treatment.

The study objective was to assess the diagnostic accuracy of clinical symptoms and their predictive valuesin patients with suspected mild COVID-19 and to identify target groups for self-isolation and outpatient treatment without additional testing in the primary health care system.

## 2. Methods

### Study design

The prospective cohort study was conducted in accordance with the Declaration of Helsinki. The study was approved by the Ethics Committee. Each study participant provided written consent to participate in the study.

### Participants

From June 2020 to December 2020, 133 persons were screened and sequentially enrolled in the study. 120 male and female outpatients aged 18 to 72 years were included in the study.

Inclusion criteria:

sudden onset of the disease,

olfactory dysfunction,

hyperthermia,

+ any of the secondary criteria: myalgia, nasal congestion, nasal discharge, cough, rhinolalia, sore throat, no signs of viral pneumonia or hypoxia, contact with a confirmed case of COVID-19, the possibility of outpatient follow-up and treatment under stay-at-home orders, signed informed consent. The inclusion criteria met the clinical criteria for mild COVID-19 in accordance with the WHO recommendations presented in the national clinical guidelines [12,23].

#### Exclusion criteria

no olfactory dysfunction, the presence of such diseases as allergic rhinitis, acute or chronic rhinosinusitis, the indications for inpatient treatment, immunodeficiency states, oncological diseases, chronic diseases of the cardiovascular or bronchopulmonary systems, diabetes mellitus, individual intolerance to the drug components.

### Test methods

Symptoms which met the diagnostic criteria for mild COVID-19 were used as indexes: hyperthermia, myalgia, nasal congestion, nasal discharge, cough, rhinolalia and sore throat. All symptoms were assessed by a physician according to a 4-point scale (0 — no symptoms, 1 — mild, 2 — moderate, 3 — severe/pronounced). Hyperthermia was assessed as: 0 (absence) < 37 °C, 1 — from 37 °C to 37.5 °C, 2 — from 37.5 °C to 38 °C, 3 — > 38 °C. In addition, patients self-assessed complaints of the severity of fever, myalgia, nasal congestion, nasal discharge, cough, rhinolalia and sore throat using a ten-point visual analogue scale (VAS). The severity of symptoms according to the VAS was assessed as: < 3 — mild, 3 to 7 — moderate, > 7 — severe.

After assessment and self-assessment of clinical symptoms, the reference standard for verification of COVID-19 — laboratory testing — was used for each patient included in the study to confirm the diagnosis. The PCR test and tests for Ig M and Ig G antibodies were carried out. The diagnosis was considered confirmed if there was at least one positive result of the above-listed tests.

ENT practitioners with experience of at least 5 years were involved in the study.

### Analysis

Diagnostic accuracy measures were assessed using the methods of descriptive statistics to describe the status of the treatment and control groups (for quantitative parameters: n, arithmetic mean, median, standard deviation, minimum and maximum values; for qualitative parameters: frequency and percentage).

The results of the inconclusive index test or reference standard were assessed using the methods of descriptive statistics to describe the status of the treatment and control groups (for quantitative parameters: n, arithmetic mean, median, standard deviation, minimum and maximum values; for qualitative parameters: frequency and percentage).

Variability analysis of the diagnostic accuracy which distinguishes the treatment group from the control group, was carried out:

A. For quantitative parameters: the normality of data distribution in groups was checked using the Shapiro — Wilk test. If the data for certain parameters in the groups were normally distributed, the groups were compared according to these parameters using Student’s t-test for independent samples (after checking the homogeneity of the variances in the groups using Levene’s test in order to choose a variant of Student’s t-test). Otherwise (the data is not normally distributed), the groups were compared using the Mann — Whitney test.
B. For categorical parameters: groups were compared using Pearson’s chi-square test. If the prerequisites for applying this criterion were not met, the Fisher’s exact test was used for comparison.

The level of significance for the Shapiro — Wilk test was taken as equal to 0.01, and for the other criteria — to 0.05.

The processing of the missing (inconclusive) data in the index test and reference standard was carried out based on the Investigators’ decisions, taking into account the effects which the inconclusive results will have, by combining the data into categories for more accurate interpretation.

Estimated sample size and method for its determination

There are no a priori estimates for the sample size in this diagnostic accuracy study. Therefore, the study included subjects in view of the economic feasibility and rule of thumb according to which a population of at least 10 patients with the target symptom was selected for each individual symptom or sign.

## RESULTS

### Participants

133 persons were screened. 120 outpatients aged 18 to 65 years were included in the study (Figure 1).

**Figure 1.**
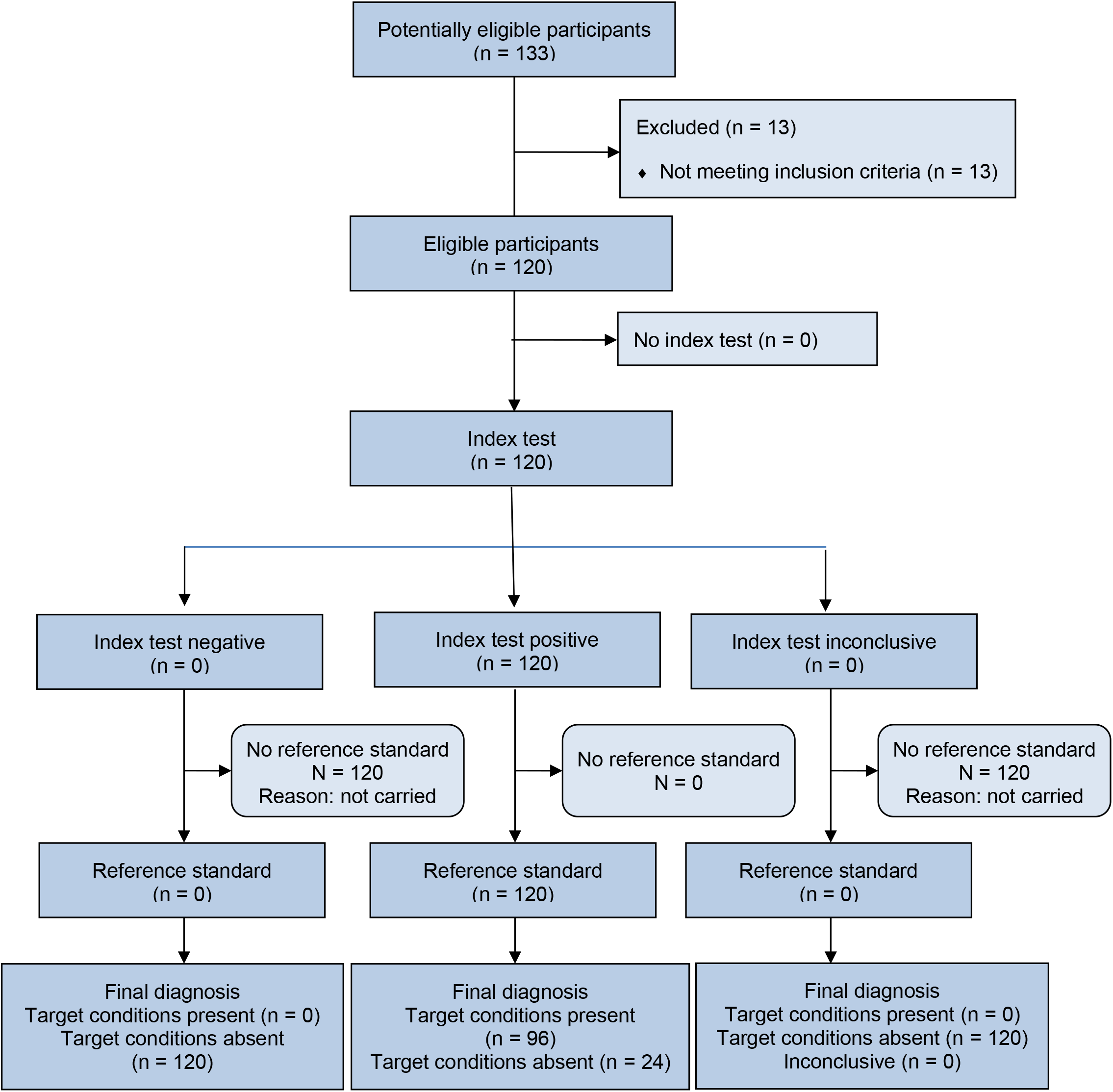
Patients included in screening and randomization

Of the 133 patients screened, 13 patients had exclusion criteria. The remaining 120 patients were randomized in the study.

Tables 1 and 2 show the distribution of patients by age and gender. The study included patients aged 18 to 72 years; the mean age was 33.44 ± 12.43 years.

**Table 1.** Allocation of patients according to age

| Parameter | Statistical parameters |  |  |  |  |  |
| --- | --- | --- | --- | --- | --- | --- |
|  | n | Arithmetical mean | Median | Standard deviation | Min | Max |
| Age | 120 | 33.44 | 30 | 12.43 | 18 | 72 |

**Table 2.** Allocation of patients according to sex

| Gender | n | % |
| --- | --- | --- |
| Male | 37 | 30.8 |
| Female | 83 | 69.2 |

### Test results

Table 3 presents an analysis of the severity of the key symptoms of mild COVID-19 in patients according to the physician’s assessment using a 4-point scale and the patient self-assessment using a 10-point VAS.

**Table 3.** Analysis of the severity of the key symptoms according to the physician’s assessment

| Parameter | Statistical parameters |  |  |  |  |  |
| --- | --- | --- | --- | --- | --- | --- |
|  | n | Arithmetical mean | Median | Standard deviation | Min | Max |
| **Physician's assessment (0–3 points)** |  |  |  |  |  |  |
| Hyperthermia | 120 | 1.84 | 2 | 1.18 | 0 | 3 |
| Myalgia | 120 | 2.50 | 3 | 1.20 | 0 | 3 |
| Nasal congestion | 120 | 1.84 | 2 | 1.30 | 0 | 3 |
| Nasal discharge | 120 | 0.89 | 1 | 1.00 | 0 | 3 |
| Cough | 120 | 0.72 | 0 | 0.99 | 0 | 3 |
| Rhinolalia | 120 | 1.18 | 1 | 1.05 | 0 | 3 |
| Sore throat | 120 | 1.25 | 1 | 1.10 | 0 | 3 |
| **Patient self-assessment (VAS 0–10 points)** |  |  |  |  |  |  |
| Fever | 120 | 3.90 | 4 | 2.82 | 0 | 10 |
| Myalgia | 120 | 5.91 | 6 | 2.92 | 0 | 10 |
| Nasal congestion | 120 | 4.08 | 4 | 3.01 | 0 | 10 |
| Nasal discharge | 120 | 1.97 | 1 | 2.47 | 0 | 10 |
| Cough | 120 | 1.49 | 0 | 2.18 | 0 | 10 |
| Rhinolalia | 120 | 2.68 | 2 | 2.59 | 0 | 10 |
| Sore throat | 120 | 3.90 | 4 | 2.82 | 0 | 10 |

The most pronounced clinical symptoms (1 point or more) were hyperthermia, myalgia, nasal congestion, rhinolalia and sore throat. Nasal discharge and cough were less pronounced (less than 1 point). According to self-assessment, fever, myalgia, nasal congestion and sore throat were assessed by the patients as more than 3 points (moderate and higher).

The diagnosis of COVID-19 was laboratory-confirmed in 96 (80%) patients out of 120 patients included in the study (Table 4).

**Table 4.**
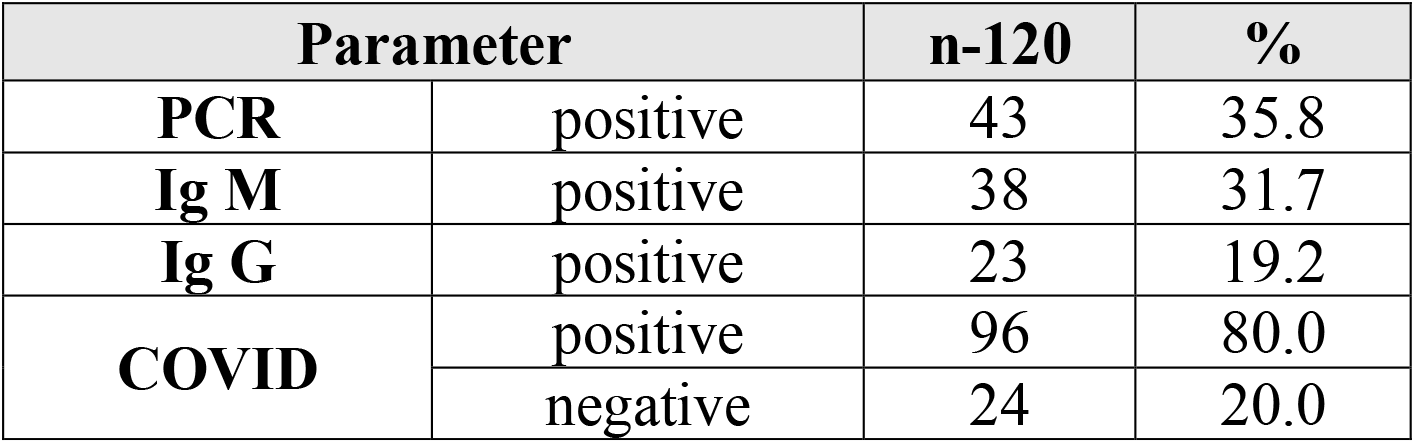
Laboratory verification analysis for COVID-19

The diagnosis was confirmed by PCR test in 43 (35.8%) patients. In the absence of verification in the first days of the disease, the patients underwent control tests (Ig M and Ig G) after the end of the self-isolation period/the end of the follow - up period. The diagnosis was verified using the method for measuring immunoglobulins in another 53 patients. In the remaining 24 (20%) patients, the diagnosis of COVID-19 was not confirmed by any of the laboratory methods.

The analysis did not show any dependence of the test results on the age of the patients. The average age of the patients with positive test results was 33.34 ± 12.00 years; with negative results — 33.83 ± 14.32 years.

We analysed the predictive values of symptoms according to the physician’s assessment and the patient self-assessment when diagnosing mild COVID-19 with descriptive statistics (Figures 2, 3).

**Figure 2.**
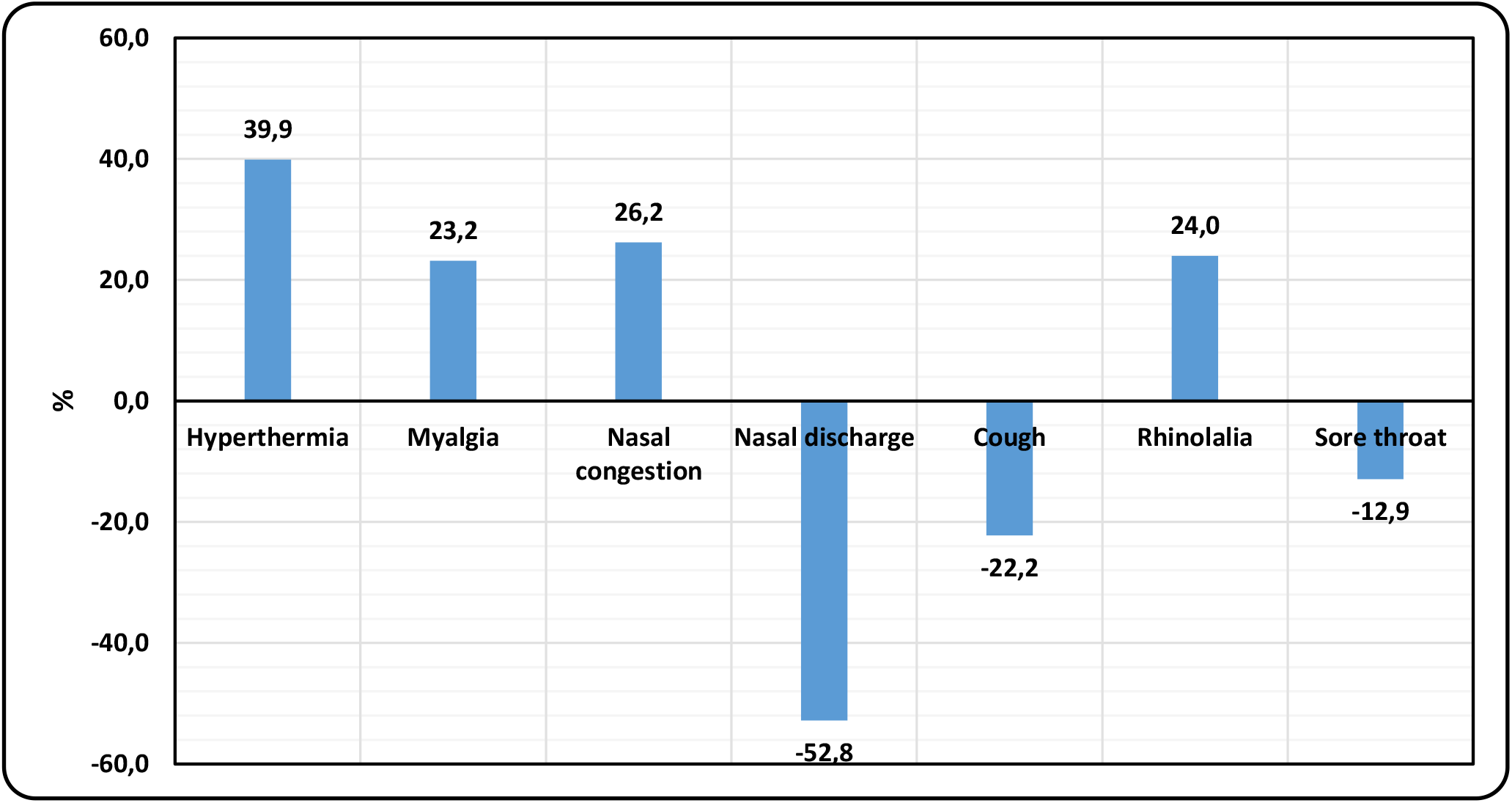
Analysis of the predictive values of the symptom severity assessed by the physician

**Figure 3.**
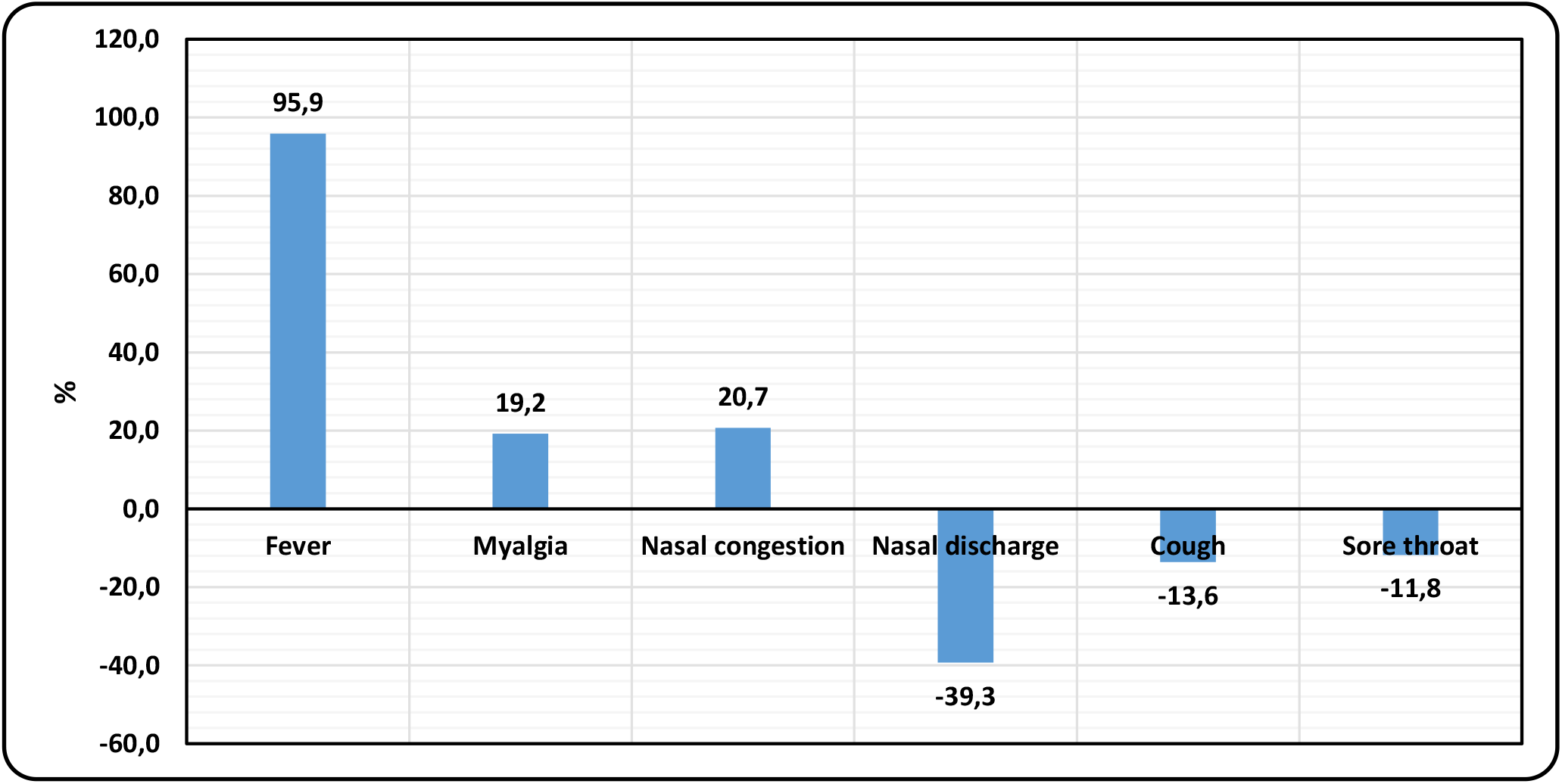
Analysis of the predictive values of the symptom severity according to patient self-assessment

As can be seen from Figure 2, according to the physician’s assessment of symptoms, the relative difference in the symptom severity in the group with a laboratory-confirmed COVID-19 diagnosis compared to an unconfirmed one is: for hyperthermia 39.0%, myalgia 23.2%, nasal congestion 26.2% and rhinolalia 24%. Nasal discharge, cough and sore throat are negative.

Thus, from the physician’s point of view, hyperthermia, myalgia, nasal congestion and rhinolalia are of great predictive significance to make a diagnosis. Nasal discharge, cough and sore throat are less significant.

According to Figure 3, the patient self-assessment of symptoms in the group with a confirmed COVID-19 diagnosis compared to an unconfirmed one, showed predictive values comparable to the physician’s assessment in the severity of fever — 95.9%, nasal congestion — 20.7%, myalgia — 19.2%. Nasal discharge, cough and sore throat have negative predictive values. Thus, from the patient’s point of view, fever, myalgia and nasal congestion are of great predictive significance to make a diagnosis. Nasal discharge, cough and sore throat are less significant.

Table 5 presents the analysis of the interaction and predictive significance of the clinical symptoms of mild COVID-19 according to the physician’s assessment and the patient self-assessment using the ordinal regression method (McCullagh method) [24].

**Table 5.** Analysis of predictive significance of symptoms using the ordinal regression method

| Model: dependent variable COVID |  | Assessment | Standard error | Wald | Degree of freedom | Significance |
| --- | --- | --- | --- | --- | --- | --- |
| **Physician assessment of symptoms** |  |  |  |  |  |  |
| Factors | Hyperthermia | 0.018 | 0.310 | 0.004 | 1 | 0.953 |
|  | Myalgia | 0.066 | 0.311 | 0.045 | 1 | 0.832 |
|  | Nasal congestion | 0.098 | 0.351 | 0.077 | 1 | 0.781 |
|  | Nasal discharge | 0.994 | 0.318 | 9.745 | 1 | 0.002 |
|  | Cough | 0.462 | 0.296 | 2.441 | 1 | 0.118 |
|  | Rhinolalia | 0.157 | 0.377 | 0.173 | 1 | 0.678 |
|  | Sore throat | 0.403 | 0.448 | 0.807 | 1 | 0.369 |
| **Patient self-assessment of symptoms** |  |  |  |  |  |  |
| Factors | Fever | 0.039 | 0.106 | 0.133 | 1 | 0.715 |
|  | Myalgia | 0.059 | 0.135 | 0.187 | 1 | 0.666 |
|  | Nasal congestion | 0.105 | 0.172 | 0.372 | 1 | 0.542 |
|  | Nasal discharge | 0.379 | 0.138 | 7.507 | 1 | 0.006 |
|  | Cough | 0.145 | 0.102 | 2.029 | 1 | 0.154 |
|  | Sore throat | 0.574 | 0.590 | 1.027 | 1 | 0.290 |
| *a. Dependent variable: COVID* |  |  |  |  |  |  |

As can be seen from Table 6, according to the physician’s assessment of symptoms, such symptoms as hyperthermia, myalgia, nasal congestion and rhinolalia with a significance level of more than 0.6 have the most pronounced predictive significance to make a clinical diagnosis of mild COVID-19. A sore throat is of low predictive significance (significance 0.36). According to the patient self-assessment of symptoms, such symptoms as fever, myalgia and nasal congestion with a significance level of more than 0.5 have the most pronounced predictive significance to make a clinical diagnosis of mild COVID-19.

Thus, taking into account the results of three methods, three factors — fever, myalgia and nasal congestion — can be distinguished which with high predictive significance have effects on making a clinical diagnosis of mild COVID-19.

## DISCUSSION

As is known, until today, there are no clinical tests specific to COVID-19 apart from laboratory confirmation. Under pandemic conditions, complete blood count, CT data, pneumonia specific to COVID and saturation values are of certain predictive value in cases of severe disease. However, none of the known symptoms are specific for making a reliable diagnosis of mild COVID-19 [13-16]. At the same time, the testing capacity under actual conditions is not enough to meet current needs. Therefore, the formation of target groups for self-isolation and outpatient treatment is an important issue [18].

The study objective was to assess the diagnostic accuracy of clinical symptoms and their predictive valuesin patients with suspected mild COVID-19, which very likely makes it possible to make a clinical diagnosis, but it has certain limitations. First, this preliminary study was retrospective with a relatively small sample size. In future, it will be needed to conduct studies with a larger sample of COVID- 19 patients. Second, the inclusion criteria included olfactory dysfunction as a mandatory marker. Therefore, it is needed to conduct future studies with a priori hypotheses regarding the role of hyposmia/anosmia in this population, since according to the literature data this symptom is not reliable, but according to our data, a number of patients with hyposmia had negative laboratory test results. Thirdly, the severity of olfactory dysfunction and its relationship with sinonasal or other symptoms remain incompletely characterized. Fourthly, all symptoms were statistically analysed only on the first day of this study without taking into account the change in condition during further follow-up. Fifthly, there were no predictive values or follow-up results for patients with disease worsening. In future, patients will be analysed over time. As a result, complex comparative studies can be conducted to study possible differences which could increase the risk of error in this study. Therefore, more data should be collected.

This study demonstrated that there were more women than men (69.2% versus 30.8%). The average age of the patients with positive test results was 33.34 ± 12.00 years; with negative results — 33.83 ± 14.32 years. According to published data, COVID-19 in males is associated with a greater disease severity and higher mortality rates [1,2]. The proportion of men with severe disease in different age groups was also higher than that of women [4]. It was also shown that age affects the outcomes of COVID-19 patients hospitalized: the elderly are more likely to have severe disease. Thus, there are significant age and gender differences between groups of patients with different disease severity, and such patients require different approaches to care, in particular, with the possibility of outpatient treatment during self-isolation even without laboratory confirmation.

An important and interesting conclusion of this study is that the intensity of these symptoms was different and depended on the physician’s assessment or the patient self-assessment. According to the physician’s assessment, the most pronounced symptoms (1 point or more) were hyperthermia, myalgia, nasal congestion, rhinolalia and sore throat; but myalgia was the most pronounced of them. Nasal discharge and cough were less pronounced (less than 1 point). According to the patient self-assessment, fever, myalgia, nasal congestion and sore throat were assessed as having more than 3 points (moderate and higher).

Another important conclusion of the study is that symptoms have different predictive effects on the diagnosis due to the relative difference in the symptom severity assessed with descriptive statistics.

According to the physician’s assessment of symptoms, hyperthermia, myalgia, nasal congestion and rhinolalia have the greatest predictive value in the group with a laboratory-confirmed COVID-19 diagnosis compared to an unconfirmed one. The relative difference in the symptom severity between groups is 24% to 39%. According to the patient self-assessment of symptoms, the difference in fever is 95%, myalgia and nasal congestion is 19.2% and 20.7%, and has a high predictive value. Nasal discharge, cough and sore throat have a negative value according to both the physician’s assessment and patient self-assessment. Due to this, their predictive diagnostic significance is low.

We analysed the interaction and predictive significance of clinical symptoms to determine the diagnosis of mild COVID-19 using the ordinal regression method. According to the physician’s assessment of symptoms, such symptoms as hyperthermia, myalgia, nasal congestion and rhinolalia with a significance level of more than 0.6 have the most pronounced effects on making a clinical diagnosis of mild COVID-19. A sore throat is of relatively low significance (significance 0.36). According to the patient self-assessment of symptoms, such symptoms as fever, myalgia and nasal congestion with a significance level of more than 0.5 have the most pronounced predictive effects on making a clinical diagnosis of mild COVID-19.

Our findings are consistent with the literature data: mild COVID-19 is characterized by well-known non-specific manifestations: fever, cough, sore throat, nasal congestion and discharge, malaise, headache and muscle pain [5, 6]. However, only qualitative characteristics (presence/absence) of these symptoms with no quantitative characteristics and assessment of effects on making a clinical diagnosis are presented in the literature.

An important and interesting conclusion of the study is that not all symptoms of mild COVID-19 listed in the clinical guidelines have the same predictive significance when making a clinical diagnosis. Such symptoms as hyperthermia, myalgia and nasal congestion are of high predictive value from both the physician’s and patient’s points of view. Rhinolalia is of high diagnostic value only when assessed by a physician. At the same time, such symptoms as nasal discharge, cough and sore throat described in the literature have almost no predictive value in making a clinical diagnosis of mild COVID-19.

Thus, hyperthermia, myalgia and nasal congestion in patients with an acute onset of the disease and the presence of olfactory dysfunction are the key symptoms prognostic of mild COVID-19. When the physician assesses the symptoms, rhinolalia is an additional key symptom of predictive value. The presence of such symptoms as nasal discharge, cough and sore throat indicates a low probability of disease. This study demonstrated that the presence of these key symptoms of predictive value can help to make a clinical diagnosis of coronavirus disease and identify target groups for self-isolation and outpatient treatment without additional testing. Highly suspect asymptomatic patients are not considered as those who have possible mild COVID- 19 infection.

## Data Availability

All the clinical trial material can be provided by corresponding author upon reasonable request

